## Supplementary Figure 1 for "Sepsis Recording in Primary Care Electronic Health Records, Linked Hospital Episodes and Mortality Records: Population-based Cohort Study in England"

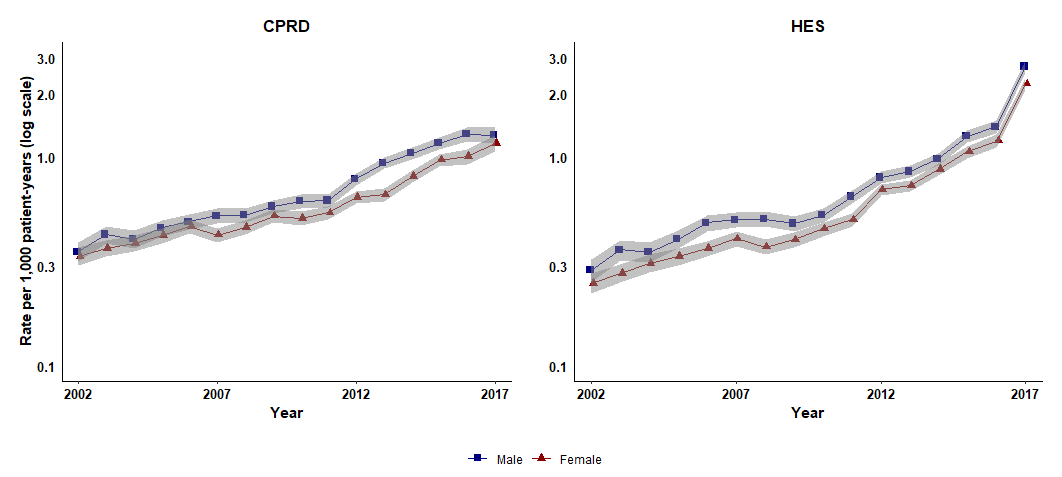

Supplementary Figure 1: Age-standardised rates of sepsis events including first events in each calendar-year.

CPRD, Clinical Practice Research Datalink; HES, Hospital Episode Statistics
