## Supplementary Figure 2 for "Sepsis Recording in Primary Care Electronic Health Records, Linked Hospital Episodes and Mortality Records: Population-based Cohort Study in England"

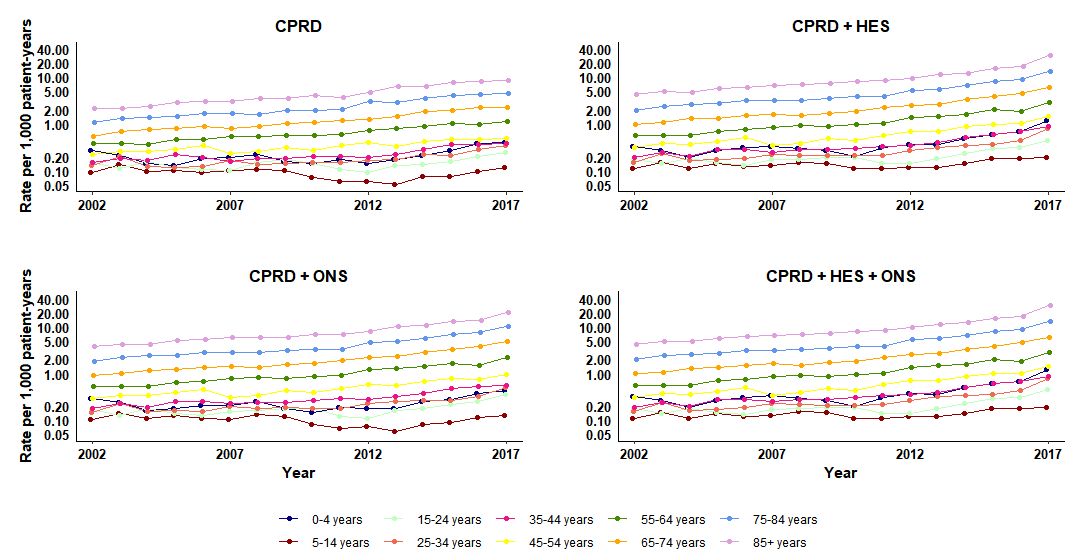

**Supplementary Figure 2: Age-specified rates of sepsis events including first events in each calendar-year.**

CPRD, Clinical Practice Research Datalink; HES, Hospital Episode Statistics; ONS, Office for National Statistics
