## Supplementary Figure 3 for "Sepsis Recording in Primary Care Electronic Health Records, Linked Hospital Episodes and Mortality Records: Population-based Cohort Study in England"

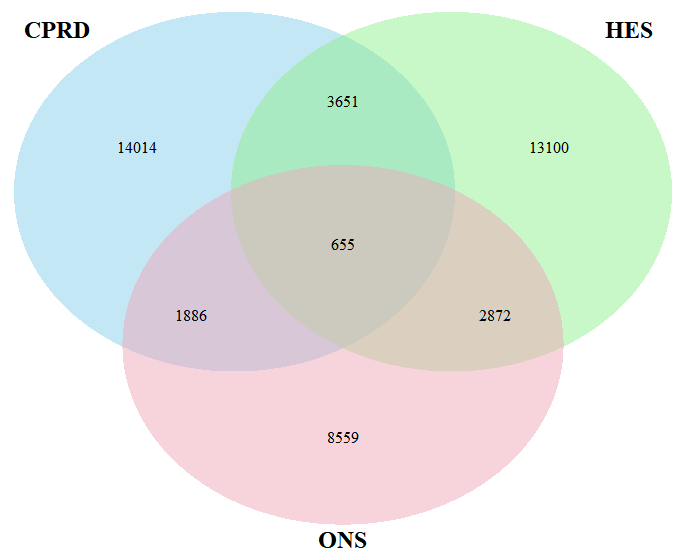

**Supplementary Figure 3: Sepsis cases in CPRD, HES, ONS (concurrent cases have index or recurrent HES sepsis events in 30 days before or after index event in CPRD or index or recurrent sepsis events within 30 days before date of death in ONS death registry).**

CPRD, Clinical Practice Research Datalink; HES, Hospital Episode Statistics; ONS, Office for National Statistics
