## Supplementary Figure 4 for "Sepsis Recording in Primary Care Electronic Health Records, Linked Hospital Episodes and Mortality Records: Population-based Cohort Study in England"

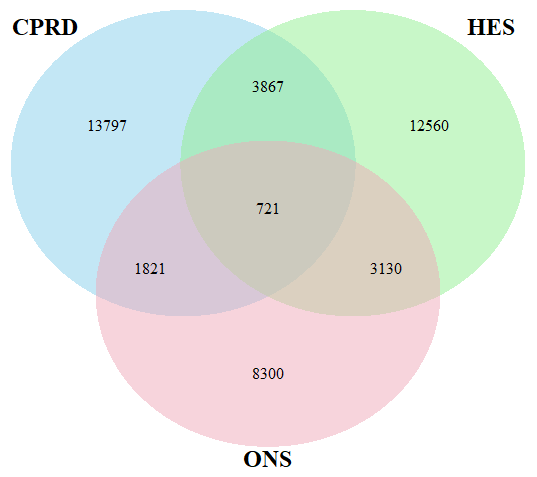

**Supplementary Figure 4: Incident first sepsis events in CPRD, HES and ONS using a 90-day time-window to evaluate concurrence.**

CPRD, Clinical Practice Research Datalink; HES, Hospital Episode Statistics; ONS, Office for National Statistics
