## Supplementary Table 1 for "Sepsis Recording in Primary Care Electronic Health Records, Linked Hospital Episodes and Mortality Records: Population-based Cohort Study in England"

**Table 1: Concurrence of sepsis events in CPRD, ONS, HES using 30-day and 90- day time-windows and either first or first and subsequent events. Figures are frequencies (% of column total).**

|  | **CPRD** | **HES** | **ONS** |
| --- | --- | --- | --- |
| **First sepsis events** | 20 206 | 20 278 | 13 972 |
| **Concurrent first sepsis event associated with first primary care sepsis record** | | | |
| 30 days | - | 4 117 (20) | 2 438 (17) |
| 90 days | - | 4 588 (23) | 2 542 (18) |
| **Concurrent first sepsis event associated with first HES sepsis record** | | | |
| 30 days | 4 117 (20) | - | 3 397 (24) |
| 90 days | 4 588 (23) | - | 3 851 (28) |
| **Concurrent first or subsequent sepsis event following first primary care record** | | | |
| 30 days | - | 4 317 (21) | 2 541 (18) |
| 90 days | - | 4 770 (24) | 2 635 (19) |
| **Concurrent first or subsequent sepsis event following first HES record** | | | |
| 30 days | 4 306 (21) | - | 3 527 (25) |
| 90 days | 4 710 (23) | - | 3 953 (28) |

CPRD, Clinical Practice Research Datalink; HES, Hospital Episode Statistics; ONS, Office for National Statistics
